# Social divisions and risk perception can drive divergent epidemic dynamics and large second and third waves

**DOI:** 10.1101/2022.05.20.22275407

**Authors:** Mallory J. Harris, Erin A. Mordecai

## Abstract

During infectious disease outbreaks, individuals may adopt protective measures like vaccination and physical distancing in response to awareness of disease burden. Prior work showed how feedback between epidemic intensity and awareness-based behavior shapes disease dynamics (e.g., producing plateaus and oscillations). These models often overlook social divisions, where population subgroups may be disproportionately impacted by a disease and more responsive to the effects of disease within their group. We hypothesize that socially divided awareness-based behavior could fundamentally alter epidemic dynamics and shift disease burden between groups.

We develop a compartmental model of disease transmission in a population split into two groups to explore the impacts of awareness separation (relatively greater in-versus out-group awareness of epidemic severity) and mixing separation (relatively greater in-versus out-group contact rates). Protective measures are adopted based on awareness of recent disease-linked mortality. Using simulations, we show that groups that are more separated in awareness have smaller differences in mortality. Fatigue-driven abandonment of protective behavior can drive additional infection waves that can even exceed the size of the initial wave, particularly if uniform awareness drives early protection in one group, leaving that group largely susceptible to future infection. Finally, vaccine or infection-acquired immunity that is more protective against transmission and mortality may indirectly lead to more infections by reducing perceived risk of infection, and thereby reducing vaccine uptake. The dynamics of awareness-driven protective behavior, including relatively greater awareness of epidemic conditions in one’s own group, can dramatically impact protective behavior uptake and the course of epidemics.

## Introduction

When an infectious disease causes substantial disease burden and death, people may respond to the true or perceived risk of infection by modifying their behavior (1–5). In turn, protective behaviors like physical distancing, mask wearing, and vaccination may suppress transmission, reducing peak and total infections and disease-linked mortality (3, 6, 7). Bidirectional feedback between epidemic outcomes and awareness-based behavior may lead to unexpected and nonlinear dynamics, such as plateaus and oscillations in cases over time (8–11). Mathematical models that split the population into categories with respect to the disease (i.e., compartments) and mathematically define transition rates between different states are widely used to understand such complex epidemic dynamics. Compartmental models may incorporate the impact of awareness as a function of deaths or cases that reduces transmission evenly across the population (8, 9). The spread of epidemic-related information has also been modeled as an additional contagion process that is distinct from but potentially linked to disease transmission (11–15). However, real populations are sharply divided in physical interactions, demography, ideology, education, housing and employment structures, and information access; these social divisions can impact both the transmission of pathogens and information within and between groups, altering epidemic dynamics. The impacts of such asymmetrically spreading disease and awareness in a highly divided population are not well understood (16–18).

Populations may be subdivided based on an array of factors (e.g., race, ethnicity, age, and geography), with marked differences in pathogen exposure and infection severity (17, 19–23). Risk of pathogen introduction may vary between groups: high income groups may encounter pathogens endemic to other regions through international travel, low income groups may have heightened likelihood of exposure connected to poor housing quality and insufficient occupational protections, and certain regions and occupations experience greater risks of exposure to zoonotic illnesses (19, 24–27). Once a pathogen is introduced, it may spread at different rates within groups based on factors like housing density and access to healthcare (20, 24, 28). Further, the severity of infection may vary directly with group identity due to underlying biological differences (e.g., age or sex), as a function of co-morbidities especially prevalent in one group due to underlying inequities (e.g., lung disease connected to environmental pollution or heart disease associated with factors driven by structural racism), or through heterogeneity in access to and quality of healthcare (20–22, 28–32). Physical barriers (e.g., geographic boundaries, schools, residential segregation, and incarceration) and preferential mixing with members of one’s own group may reduce contact and subsequent transmission between groups, a characteristic we describe as separated mixing (19, 33–36). Infectious disease models that account for differences in vulnerability within subgroups of a population and separated mixing can help to illustrate the emergence of health inequities and justify structural interventions to reduce these disparities (37–40). However, such models may miss an important behavioral dimension by failing to account for variation in awareness-based behavior changes among groups.

Awareness and behavioral heterogeneity can significantly alter disease dynamics: for example, local awareness in a network with strong clustering can stop the pathogen from spreading altogether, while clustering in vaccine exemptions may lead to outbreaks (14, 41, 42). Although personal risk perception may be responsive to risk in other groups, and behavior may be influenced by population-level social norms and mass media, attitudes toward diseases and protective behaviors may also vary considerably between groups and correspond to actual risk and personal experiences of close social ties with the disease (43–48). While prior awareness-based models have examined outcomes given different sources of information (i.e., local or global), we aim to characterize risk perception based on group-level information in a population split into two distinct and well-defined groups (49). We define separated awareness as greater in-versus out-group awareness in a split population and predict that, by producing behavioral responses more reflective of each group’s risk, it may reduce differences between groups in disease burden (50). Understanding the impacts of separation with respect to mixing and awareness on disease dynamics may be important for characterizing differences in epidemic burden and effectively intervening to mitigate population inequities (37, 39, 40, 50, 51).

Here, we investigate the impacts of intergroup divisions on epidemic dynamics using an awareness-based model for transmission of an infectious disease, in which adoption of protective measures (either nonpharmaceutical interventions or vaccinations) is linked to recent epidemic conditions and mediated by awareness.

We ask:

1. How do separated awareness and mixing interact to affect differences between groups in epidemic dynamics?
2. How does fatigue interact with awareness separation to affect long-term epidemic dynamics?
3. When vaccines are introduced, how does immunity interact with awareness separation to affect long-term epidemic dynamics?

## Results

### 1. Separated mixing and awareness

To understand how separation in awareness and mixing interact to alter short-term epidemic dynamics in a split population, we model awareness-based adoption of nonpharmaceutical interventions (Equation 1); all model parameters are defined in Supplementary Table 1 and a compartmental diagram is provided as Supplementary Figure 1. The population is split into two groups: group *a* and group *b*, and individuals in each group can switch between unprotective behavior and protective behavior that reduces transmission but cannot change their group. We arbitrarily designate group a as having greater underlying vulnerability to infection or disease-linked mortality in all of the following scenarios. Specifically, in this section the sole initial difference between groups is caused by introducing the pathogen into group *a* alone at prevalence *I*_*a*_ (0) = 0.001; all other parameters are equivalent between groups. To simplify short-term wareness-based behavior, this scenario does not incorporate memory or fatigue (ℓ and *ϕ* = 0,). First, we allow both mixing (*h*; which drives the contact and contagion uniform (functioning like a single population) or highly separate(Prosses). process and awareness (*ϵ* which drives protective behavior adoption) to be either uniform (functioning like a single population) (0.5) or highly separated (0.99).

The groups experience identical epidemic dynamics when mixing is uniform (Figure 1A, B), as the pathogen introduced into group *a* quickly spreads into group *b* and circulates evenly within and between groups. When groups mix separately, differences in epidemic dynamics between groups arise and depend on awareness separation (Figure 1C, D). When mixing is separated but awareness is uniform, epidemic shape differs in both timing and magnitude between groups, increasing the peak size and total infections in the more vulnerable (earlier epidemic introduction) group *a* and decreasing both in group *b* (Figure 1C). Specifically, uniform awareness reduces total infections in group *b*, which adopts protective behavior by observing mortality in group *a* at a point when infections within group *b* remain relatively low (Figure 1C, Supplementary Figure 3B, D, E). Meanwhile, uniform awareness causes group *a* to underestimate disease severity due to the lack of early mortality in group *b*, leading to decreased early protective behavior and a larger outbreak (Figure 1C, Supplementary Figure 3A, C, E). When awareness is separated, group *b* has little awareness of the emerging epidemic localized to group *a*, while group *a* responds to its relatively higher early disease burden with increased awareness, driving epidemic dynamics between the two groups to be similar in shape but delayed in time for group *b* (Figure 1D). Therefore, awareness separation reduces the differences between groups in epidemic shape (e.g., peak size, total infections), while mixing separation offsets them in time (Figure 1C, D).

**Figure 1.**
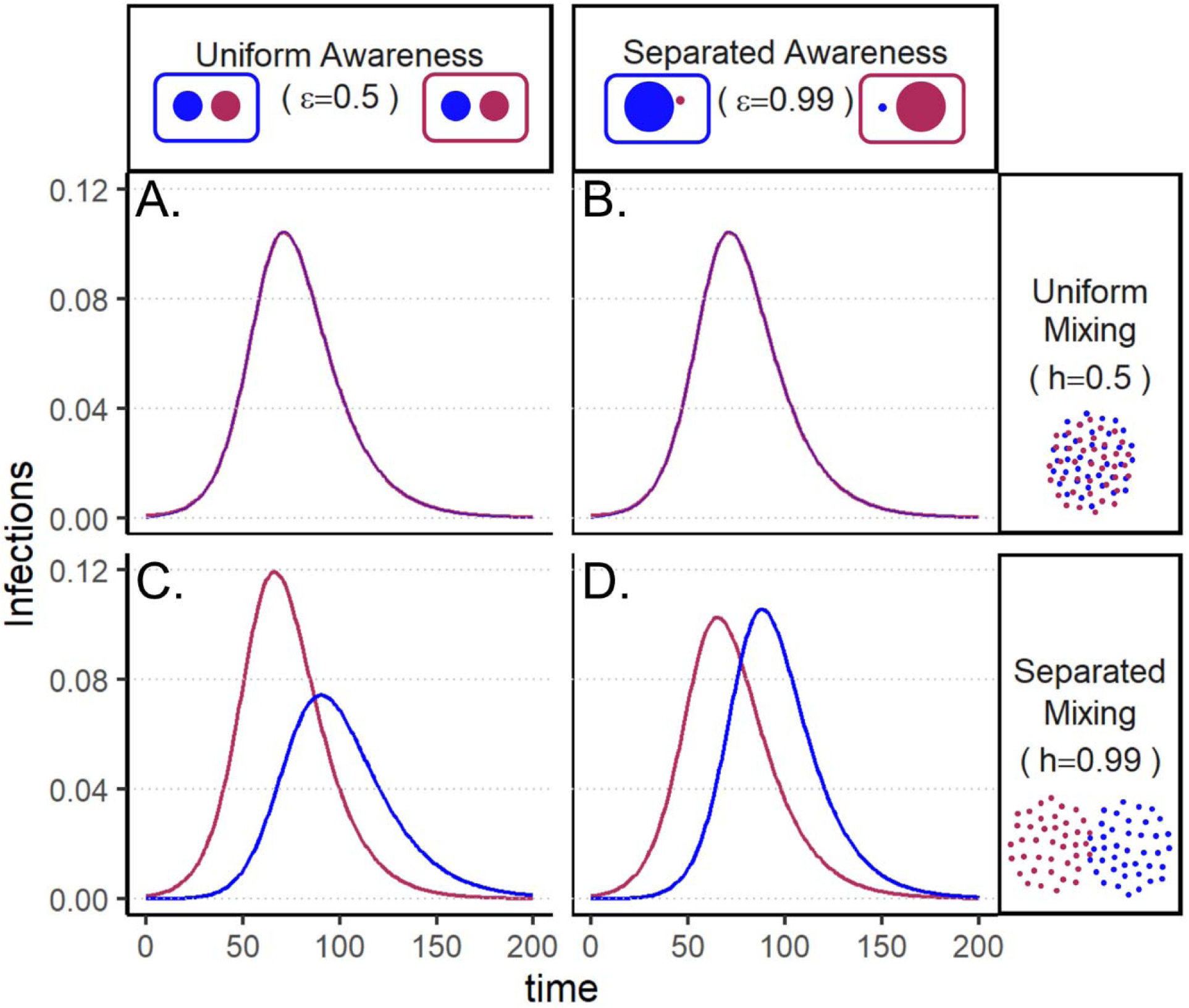
Epidemic peaks are offset in time between groups when mixing is separated (C, D), and in magnitude when awareness is uniform but mixing is separated (C). Plots show numbers of infections over time in group a (maroon) and group b (blue) under four scenarios: awareness is uniform (A, C;*ϵ* = 0.5) or separated (B, D; *ϵ* = 0.99); mixing is uniform (A, B; *h* = 0.5) or separated (C, D; *h* = 0.99). We assume the pathogen is introduced only in group a (maroon) at prevalence 0.001 and that all other parameters are equivalent between groups: transmission coefficient (*β* = 0.2), infectious period 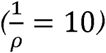, infection fatality rate (*µ* = 0.01), protective measure efficacy (*k* = 0.3), responsiveness (*θ* = 100), memory (*ℓ* = 1), and fatigue (*ϕ*= 0). Lines overlap under separated mixing (top row).

The finding that awareness separation reduces differences between groups in severe outcomes also holds when groups differ in their transmission coefficients, infectious periods, and infection fatality rates (Supplementary Figures 4, 5, 6).

### 2. Fatigue and awareness separation

We introduce memory and fatigue to examine the long-term impacts of separated awareness when awareness-driven protective behavior is abandoned over time. Once again, the pathogen is introduced into group *a* alone and all other parameters are equivalent between groups. To maintain between-group differences, we assume separated mixing (h = 0.99).

In all cases, when protective behavior wanes with fatigue, three distinct peaks emerge before transmission plateaus at low levels and declines gradually (Figure 2). The initial difference between groups with uniform awareness (Figures 1C, 2A) means that group *b* retains a relatively larger proportion of susceptible individuals who avoided infection in the first wave by rapidly adopting protective behaviors. As a result, the second and third wave in group *b* exceed its first wave in peak and total infections (Figure 2A). Meanwhile, uniform awareness causes the second wave in group *a* to be smaller and delayed by about 400 days compared to separated awareness (Figure 2A vs. B). As shown in the case without memory and fatigue (Figure 1), when both mixing and awareness are separated, the groups differ mainly in the timing of epidemic peaks rather than in their magnitude, before converging on a long and slow decline (i.e., shoulder; Figure 2B) (9).

**Figure 2.**
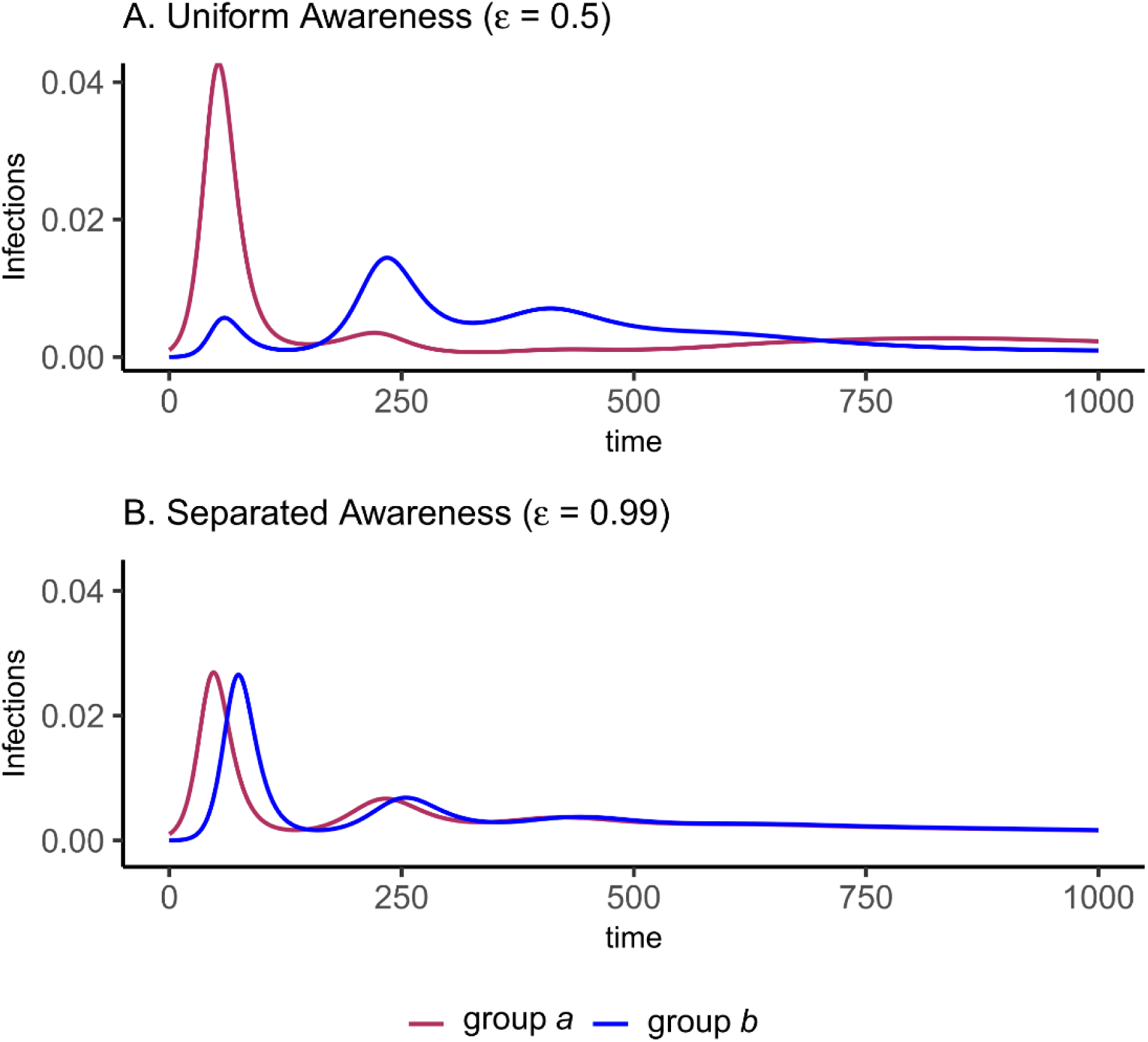
Fatigue and long-term memory produce multiple epidemic peaks, which exceed the size of the initial peak in group b when uniform awareness and separated mixing leave that group with a high proportion of susceptible people following the first wave. We initialize the model with separated mixing (*h* = 0.99), long-term memory (*ℓ* = 0.5), and fatigue (*ϕ* = 0.02); all other parameters are the same as in Figure 1. We consider infections in group a (maroon) and group b (blue) over a longer time period (1000 days, compared to 200 in Figure 1). The panels correspond to (A) uniform awareness (*ϵ* = 0.5) and (B) separated days awareness (*ϵ* = 0.99).

### 3. Immunity and awareness separation

Next, we consider the implications of awareness-based vaccine uptake in a split population given waning immune protection against infection and durable protection against mortality (Equation 3, Supplementary Figure 2). We model immunity from prior infection as equivalent to immunity from vaccination. Unlike in the previous analyses, the pathogen is now introduced at the same prevalence in both populations simultaneously to ensure that group *a* and *b* begin the post-vaccine period with similar levels of immunity. Group differences are driven by an infection fatality rate in group *a* that is twice that of group *b*. Again, we assume separated mixing (h = 0.99) to maintain distinct dynamics between the groups.

After an initial large wave, vaccination and waning immunity lead to damped cycles of infections and deaths (Figure 3). As was the case with the nonpharmaceutical intervention model (Figure 1), when awareness drives vaccination behavior, separated awareness helps to reduce differences between-group differences in mortality (Figure 3D vs. C). Group *a* becomes vaccinated at a higher rate in response to the greater number of deaths observed in group *a*, an effect that is most notable during the second epidemic peak (Figure 3D). Therefore, with separated awareness group *a* also has fewer infections than group *b* in later waves (Figure 3B), while infection dynamics remain identical (despite the larger disparity in deaths) in the uninform awareness scenario (Figure 3A), the opposite of the nonpharmaceutical intervention scenario (Figure 2).

**Figure 3.**
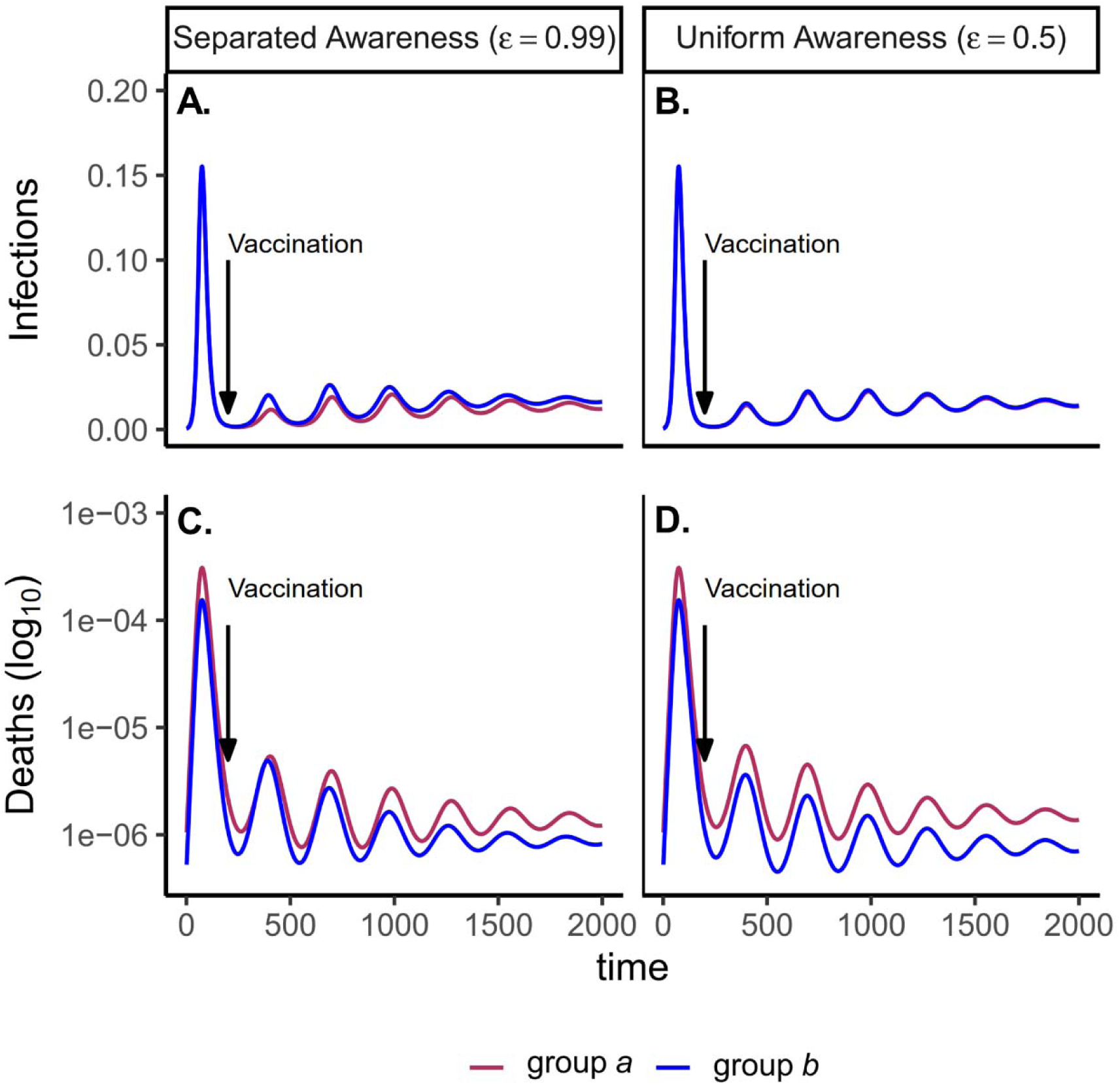
Waning immunity and awareness-based vaccination drive epidemic cycles; separated awareness reduces the disparity in deaths (C vs. D) as more-vulnerable group a members become vaccinated at a higher rate. We consider infections (A, B) and deaths (C, D) given awareness-based vaccination, where vaccination begins at day 200, indicated with vertical arrows. In the pre-vaccine period, regardless of awareness separation, infection dynamics to a doubly high infection fatality rate (*µ*_*a*_ = 0.02 and *µ*_*b*_ = 0.01; C, D). In the post-vaccine are identical between groups but deaths are higher in group a (maroon) than group b (blue) due period, we compare uniform awareness (*ϵ* = 0.5) (A, C) and separated awareness (*β* = 0.2) (B, D). Other parameter values are: *k* = 0.05 (transmission coefficient), *ζ* = 0.5 (transmission-reducing immunity), ω = ϕ = 0.5 (mortality-reducing immunity), (waning immunity), infectious period 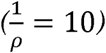 (responsiveness), ℓ = 30 (memory), *h* = 0.99 (separated mixing), *I*_0_ = 0.001 (initial infection prevalence).

Because vaccination protects against infections and deaths, and recent deaths feed back to influence awareness-driven vaccine uptake, we explored the tradeoff between immune protection and epidemic dynamics in the post-vaccine period. Assuming that vaccination and infection reduce both the transmission coefficient and infection fatality rate to an equivalent extent, we examine the total effect of variation in immune protection on epidemic dynamics and their feedbacks on vaccine uptake rate. As expected, greater immune protection reduces the number of deaths by directly reducing the infection fatality rate. However, because of awareness-driven vaccine uptake, vaccination can produce diminishing returns at the population scale where doubling immune protection from death and infection only reduces total deaths by about one eighth due to the compensatory reduction in vaccine uptake (Figure 4A), despite doubling individual protection for vaccinated people. Since a more effective immune response reduces mortality, the perceived risk associated with infection declines and fewer people become vaccinated (Figure 4B). The tradeoff between the direct impacts of immune protection on preventing infections and reduced uptake produces a nonlinear relationship between total infections and immune protection (Figure 4C). At low immune protection, infections remain approximately constant as immune protection improves. At higher levels of immune protection, reduced uptake with improving immune protection leads to more infections (Figure 4C).

**Figure 4.**
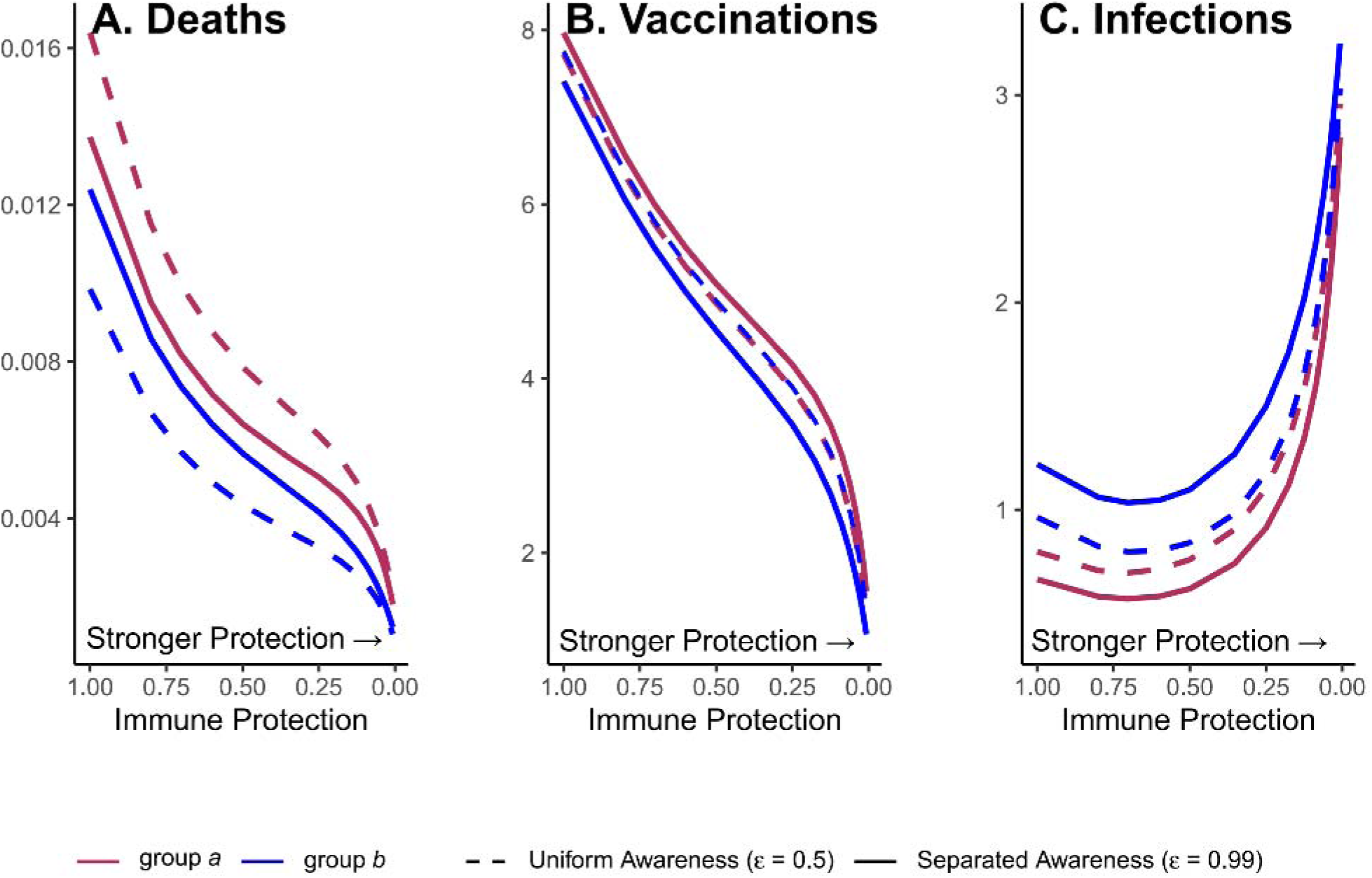
Greater immune protection (from vaccination and infection) leads to lower death rates (A), which in turn decreases vaccination rates (B) and increases infection rates (C); separated awareness reduces disparities in death rates (A) as groups are vaccinated at different rates proportional to their risks of death (B), creating differences in infection rates (C). We vary transmission-reducing immunity and mortality reducing immunity, assigning both parameters the same values (*k* = *ζ*) and define this quantity as immune protection, which we assume is equivalent for vaccine- and infection-derived immunity. The x-axis is reversed because smaller values indicate stronger protection. We examine the impacts of stronger immune protection (lower values of *k* and ζ) on total deaths (A), vaccinations (B), and infections (C) in the post-vaccine period (t = 200 through t = 2000), depending on awareness separation. We compute each quantity for group a (maroon) and group b (blue) given uniform (dashed lines ; *ϵ* = 0.5 or separated (solid lines; *ϵ* = 0.99 awareness. Other parameter values are the same as Figure 3.

Separated awareness drives greater differences between groups in vaccination behavior—the higher-risk group *a* gets vaccinated at a higher rate in response to awareness of the higher numbers of deaths in that group (Figure 4B). This in turn increases differences in infections (group *a* experiences lower infection rates; Figure 4C) but decreases differences in mortality between groups (death rates are lower for group *a* but higher for group *b* than in the uniform awareness scenario; Figure 4A). Since group *a* is at a higher inherent risk of mortality given infection, separated awareness differentially promotes vaccination and reduces infection in this group, while uniform awareness misleads group *a* into ignoring its higher risk of mortality (Figure 4A, B, solid versus dashed lines).

## Discussion

Awareness separation and social divisions may interact to fundamentally alter disease dynamics, creating or erasing differences among groups in the timing and magnitude of epidemic peaks. Uniform awareness can exacerbate differences between population subgroups when the more vulnerable group (e.g., the group where the pathogen is introduced or the group with higher infection fatality rates) underestimates the in-group risk of disease and fails to adopt early protective measures (Figures 1, 4). At the same time, the initially less-vulnerable group receives indirect protection from observing and responding to epidemic effects in the more vulnerable group, adopting protective measures that reduce their total and peak infections (Figures 1, 4). However, when awareness-driven behavior fades with fatigue, the relative disease burden may shift between groups such that the group that initially had fewer infections has relatively more infections in subsequent waves, especially when uniform awareness protects the initially less-vulnerable group during the first wave of infection (Figure 2). Awareness separation diminishes between-group differences in severe outcomes (Figures 1, 2, 3, 4, Supplementary Figures 3, 4, 5, 6), but may do so by increasing differences in behavior and infections (Figures 3, 4, Supplementary Figure 6). For example, when the more vulnerable group has a higher rate of disease-linked mortality, awareness separation leads them to have higher vaccine uptake in response to their heightened perceived (and actual) risk, narrowing the difference in mortality (Figure 4). More broadly, awareness separation generally closes differences between groups by producing preferential uptake of preventative measures by the group with the greatest recent mortality, which is usually the group at greatest current risk.

In this model, greater awareness separation generally reduces differences in severe outcomes between groups because the awareness process explicitly responds to severe outcomes (deaths). But the magnitude of these impacts may vary depending on behavioral and social processes. To assess the robustness of our conclusions about the effects of awareness separation, the same scenarios could be evaluated across different models of awareness-based behavior changes, including saturation at a certain threshold for deaths (9), consideration of both lethal and non-lethal impacts of disease (e.g., hospitalizations and cases), or optimization to balance the benefits of protection against the costs of various measures (8, 10, 52). The latter approach may clarify a point that is not addressed in our analysis: although awareness separation may reduce disparities in severe disease-linked outcomes, this phenomenon is not necessarily equitable or desirable. In fact, if self-protection is associated with significant costs, already-vulnerable populations may suffer compounding costs as they balance self-protection against significant disease risk without adequate support from a broader community that does not share their risks (52–55). Further, structural inequities often leave population subgroups that are vulnerable to larger, more severe outbreaks with reduced access to protective measures like health education, treatment, vaccination, and paid leave (5, 20, 46, 48, 56–60). Resulting differences in rates of protective behavior uptake and effectiveness can compound disparities between groups and reduce the protective impact of awareness separation for more-vulnerable groups.

Epidemics are complex phenomena that typically involve heterogeneous mixing among groups of people that differ in biological and social risk factors, dynamic evolution of host behavior, pathogen infectiousness, and immune evasion, and ever-changing epidemiological and policy responses to real and perceived risk. Despite this range of potential drivers, we show here that a simple model that captures two key social processes—awareness-driven protective behavior in a split population that can be separated in mixing and awareness—can drive many of the complex dynamics observed in emerging epidemics like Covid-19. For example, when awareness is uniform and mixing is separated, the group in which the pathogen is introduced later can experience second and third waves that exceed the initial wave in size (Figure 2). This trend resembles one observed in the United States during the first year of the Covid-19 pandemic, where certain regions where the virus was introduced early (e.g., New York City metropolitan area) experienced a large early wave and relatively few infections over the rest of the year, while other regions (e.g., the southern United States) generally had small early waves and larger second and third waves. Many hypotheses have been introduced to explain this phenomenon (e.g., seasonal climate factors and population density) and several factors may have contributed to this pattern (61, 62). Yet, in our model these dramatic differences among populations in epidemic waves occur despite the groups being identical in transmission rates and disease outcomes and are entirely due to awareness-driven behavior with uniform awareness among groups (Figure 2). Although the current analysis does not examine causation, we have demonstrated how a simple behavioral process can qualitatively reproduce complex epidemic dynamics observed in real populations.

Feedback between vaccine efficacy and awareness-based vaccine uptake can also produce the counterintuitive scenario where vaccines that cause a greater reduction in transmission and mortality lead to more total infections, even as deaths are reduced (Figure 4). If, as we assume here, protective behavior is driven by awareness of severe outcomes like mortality, awareness separation may reduce differences in deaths between groups while widening differences in cases (Figures 3, 4). Accounting for awareness-based adoption of protective behavior is therefore critical for understanding complicated epidemic dynamics such as plateaus and cycles (Figures 2, 3), accurately deploying protective measures, and assessing their impact across different diseases and population subgroups (8, 9, 50).

Here we have considered arbitrarily defined groups that can be separated in mixing and awareness but initially differ only in the timing of pathogen introduction (Figures 1, 2) or in infection fatality rate (Figures 3, 4), but real social groupings may fall along a number of social, demographic, and geographic lines. The most relevant groupings with respect to awareness and disease risk may depend on the disease, while the assumption of two distinct and identifiable groups may not fully capture relevant social dynamics. For infectious diseases that are generally more prevalent and severe in children (e.g., pertussis and measles), risk may depend on age while awareness is split between parents of young children versus adults without children or among parents with different sentiments towards childhood vaccination (63). In the context of Covid-19, disease burden and attitudes toward preventative measures (e.g., masks and vaccines) have differed markedly across race, age, and socioeconomic status and over time, demonstrating how intersecting and imperfectly overlapping identities may interact to determine attitudes, protective behaviors, and risk (64–66). Moreover, ideological and social factors that do not correspond directly to disease risk (e.g., political affiliation) may influence decision-making and cause the level of protective behavior in certain subgroups to diverge sharply from their relative risk for severe disease, potentially overcoming the effects of awareness separation (46, 67). This process could be incorporated into our model by splitting the population into additional groups with respect to a cultural contagion or (mis)information spread process and allowing protective measures to be adopted based on awareness or contact with protective in-group members and rejected through fatigue or aversion to protective measures displayed by the opposite group (68, 69).

Although we assumed that awareness was directly proportional to recent mortality, external influences like partisanship (46, 67), media coverage (70), misinformation (71), and policy (3) may alter the perception of risk or the adoption of protective measures at both the individual and group level. Group identification and assessment of relative risk may be unclear or inaccurate based on uncertainty at the beginning of the outbreak, misinformation about risk factors, a gradient in risk (e.g., gradually increasing risk with age), lack of data stratification, or unobserved risk factors. Attitudes based on one disease may carry over to another disease even if risk factors differ. Relative risk across groups may also vary across time and space, potentially leading to inaccurate assessment based on prior conditions: for example, a mild initial epidemic wave can mislead a group into believing they are inherently more protected and thereby relaxing protective behaviors. Cognitive interventions that increase the accuracy of individual risk perception, especially in high-risk groups, may help to reduce between-group differences in disease burden (72, 73). To realistically capture actual behavioral responses to disease outbreaks and to understand the extent of awareness separation in real populations, our model could be parameterized using a combination of epidemiological, survey, mobility, and social media data (9, 74, 75). Considering awareness separation as a social process that may interact with mixing, fatigue, waning immunity, pathogen evolution, and pharmaceutical and non-pharmaceutical interventions may help to explain how humans are affected by and respond to infectious diseases in the presence of social divisions.

## Methods

### Nonpharmaceutical intervention model

We model disease transmission with awareness-based adoption of nonpharmaceutical interventions that reduce transmission rates. See Supplementary Figure 1 for a compartmental diagram for this model and Supplementary Table 1 for parameter definitions. We model disease transmission with a Susceptible-Infectious-Recovered-Deceased (SIRD) model, tracking the proportion of the population in each compartment through time. New infections arise through contact between susceptible and infected individuals, with transmission coefficient *β*. Individuals exit the infectious compartment at per capita rate *ρ*, the inverse of infectious period 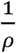 and either recover or die. The infection fatality rate, or fraction of individual exiting the infectious compartment who die, is *µ* (meaning that recovery after infection occurs with probability 1-*µ*).

We further categorize the population based on whether they adopt behavior that is Protective (P) or Unprotective (U). Compartment names contain two letters, the first indicating disease status and the second indicating behavior (e.g., SU denotes Susceptible people with Unprotective behaviors). We track the attitudes of Recovered and Deceased individuals (at the time of death), although they do not contribute directly to transmission. Protective measure efficacy against infection is indicated by a scaling factor *k* (where *k* = 0 corresponds to complete protection and *k* = 1 corresponds to no protection). Protective measures affect the behavior of both susceptible and infected individuals, so transmission rate is reduced by a factor of *k*^2^ in encounters where both parties have adopted protective measures. Living individuals can switch between protective and unprotective attitudes. Unprotective individuals adopt protective behaviors based on awareness (*α* (*t*)), which is the product of deaths over the past *ℓ* days (making *ℓ* a measure of memory) and a responsiveness constant *θ*. Protective behaviors are abandoned due to fatigue at per capita rate *ϕ*.

The population is split into two groups of equal size, where group membership is fixed, and each group contains all epidemiological compartments. The groups are labelled as *a* and *b* and indicated as a subscript in compartment names (e.g., *SU*_*a*_ corresponds to Susceptible-Unprotective individuals in group *a*). Parameters may vary between groups, as indicated by subscripts (e.g *θ*_*a*_ corresponds to responsiveness in group *a*).If parameters are equivalent for both groups, we exclude the subscript (e.g., *θ* = *θ*_*a*_ = *θ*_*b*_).

Preferential within-group mixing is represented by homophily parameter *h*, corresponding to the proportion of contacts that are within-group. When *h* is 0.5, mixing is *uniform*, meaning that individuals are equally likely to contact members of their own group as members of the opposite group. As *h* approaches 1, mixing becomes increasingly separated, meaning that contacts are increasingly concentrated within groups. Similarly, we consider separation in awareness, *ϵ*, or the relative weight of in group versus out-group awareness of deaths for protective behavior.

The system of equations for group *a* is as follows (equations for group *b* can be derived symmetrically):

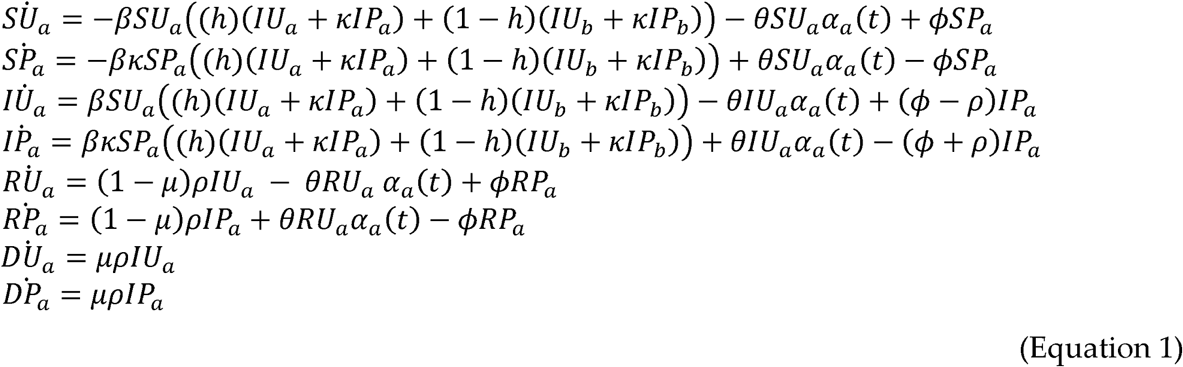

Where *α*_*a*_ (*t*) the awareness equation for group *a*:

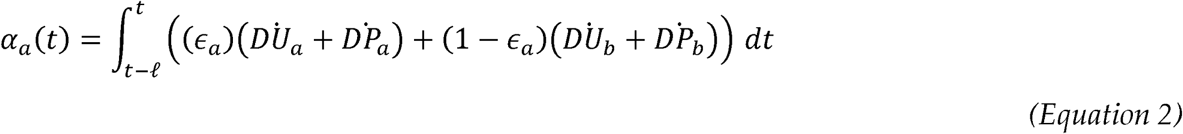

### Vaccination model

We develop an alternative model of awareness-based vaccine uptake. See Supplementary Figure 2 for a compartmental diagram for this model and Supplementary Table 1 for parameter definitions. Here, the second letter of compartment names indicates immune status: Unprotective (U), Transmission and Mortality-Reducing Immunity (T), or Mortality-Reducing Immunity (M).

As in the nonpharmaceutical intervention model, susceptible people without prior immunity (SU) may become infected and then recover or die according to baseline infection parameter values. Susceptible individuals may become vaccinated and transition directly to the recovered compartment, bypassing infection, at a rate dependent on the awareness equation (Equation 2). There may be a lag between the beginning of the epidemic and vaccine introduction at time point *t*_*v*_. To evaluate long-term immune effects of vaccination and infection on epidemic dynamics, we incorporate waning immunity.

After vaccination or infection, individuals temporarily have complete protection from infection (RT). At per capita rate *ω*, they regain susceptibility to infection, this time with transmission and mortality-reducing immunity (i.e., *ST*). As in the nonpharmaceutical intervention model, transmission-reducing protection scales transmission rates for susceptible and infected individuals by a constant. Additionally, immunity from infection reduces disease-linked mortality by scaling factor *ζ*. Transmission-reducing immunity is lost at per capita rate *ϕ*, while the ortality-reducing immunity is retained over the course of the simulation, reflecting how neutralizing antibody production may decay over time while cellular immune responses are more durable (76). Susceptible individuals with mortality-reducing immunity alone (*SM*) may regain transmission-reducing immunity via vaccination, which occurs based on the same awareness function as vaccination of people without immune protection.

The system of equations for this model in a population without groups is:

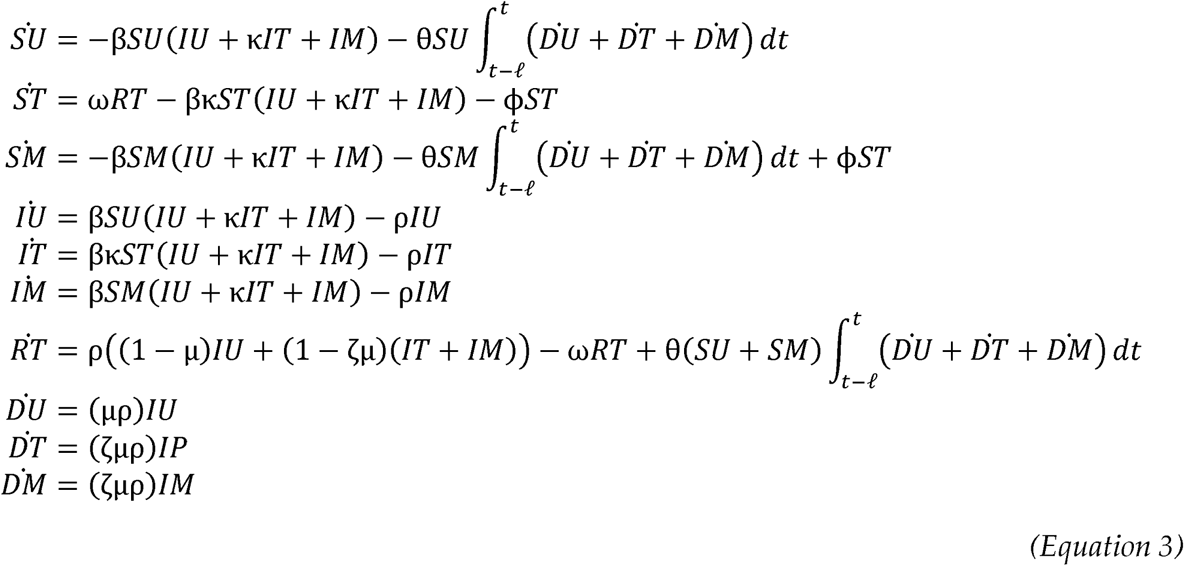

The equations for a split population with separated mixing and awareness can be derived following Equation 1.

### Simulations

We ran simulations in R version 4.0.2, using the dede function in the deSolve package, which solves systems of differential equations (77). The population begins as almost fully susceptible (*s* (0)≈ 1), with a small initial infection prevalence (*I* (0)) to seed the outbreak and no protective behaviors.

## Data Availability

Code used to conduct these analyses are available on Github at: https://github.com/mjharris95/divided-disease

## Author contributions

MJH and EAM conceived of project and designed models; MJH conducted analyses; EAM and MJH interpreted simulations and wrote the manuscript.

## Supplementary files

**Supplementary Table 1.** Parameter dictionary providing parameter symbols, descriptions, and values for different scenarios. The parenthetical numbers in the values column indicate the scenario where the parameter takes the given values (1: separated mixing and awareness; 2: fatigue and awareness separation; 3: immunity and awareness separation).

| Parameter | Description | Value |
| --- | --- | --- |
| $\beta$ | Transmission coefficient | 0.2 (1, 2, 3) |
| $1/\rho$ | Infectious period | 10 (1, 2, 3) |
| $\mu$ | Infection fatality rate | 0.01 (1,2);<br>$\mu_a = 0.01$ and $\mu_b = 0.02$ (3) |
| $\kappa$ | Transmission and infection reduction with protective behavior | 0.3 (1, 2); 0.05 (3) |
| $\theta$ | Responsiveness | 100 (1, 2); 20 (3) |
| $\ell$ | Memory | 0 (1); 30 (2, 3) |
| $\phi$ | Fatigue (NPI model)/waning transmission-reducing immunity (vaccine model) | 0 (1); 0.02 (2); 0.01 (3) |
| $h$ | Homophily | 0.5 (uniform) or 0.99 (separated) (1);<br>0.99 (separated) (2, 3) |
| $\epsilon$ | Assortative awareness level | 0.5 (uniform) or 0.99 (separated) (1, 2, 3) |
| $\omega$ | Return to susceptibility (waning immunity) | 0.01 (3) |
| $\zeta$ | Mortality reduction | 0.05 (3) |
| $t_v$ | Vaccination Start Time | 200 (3) |

**Supplementary Figure 1.**
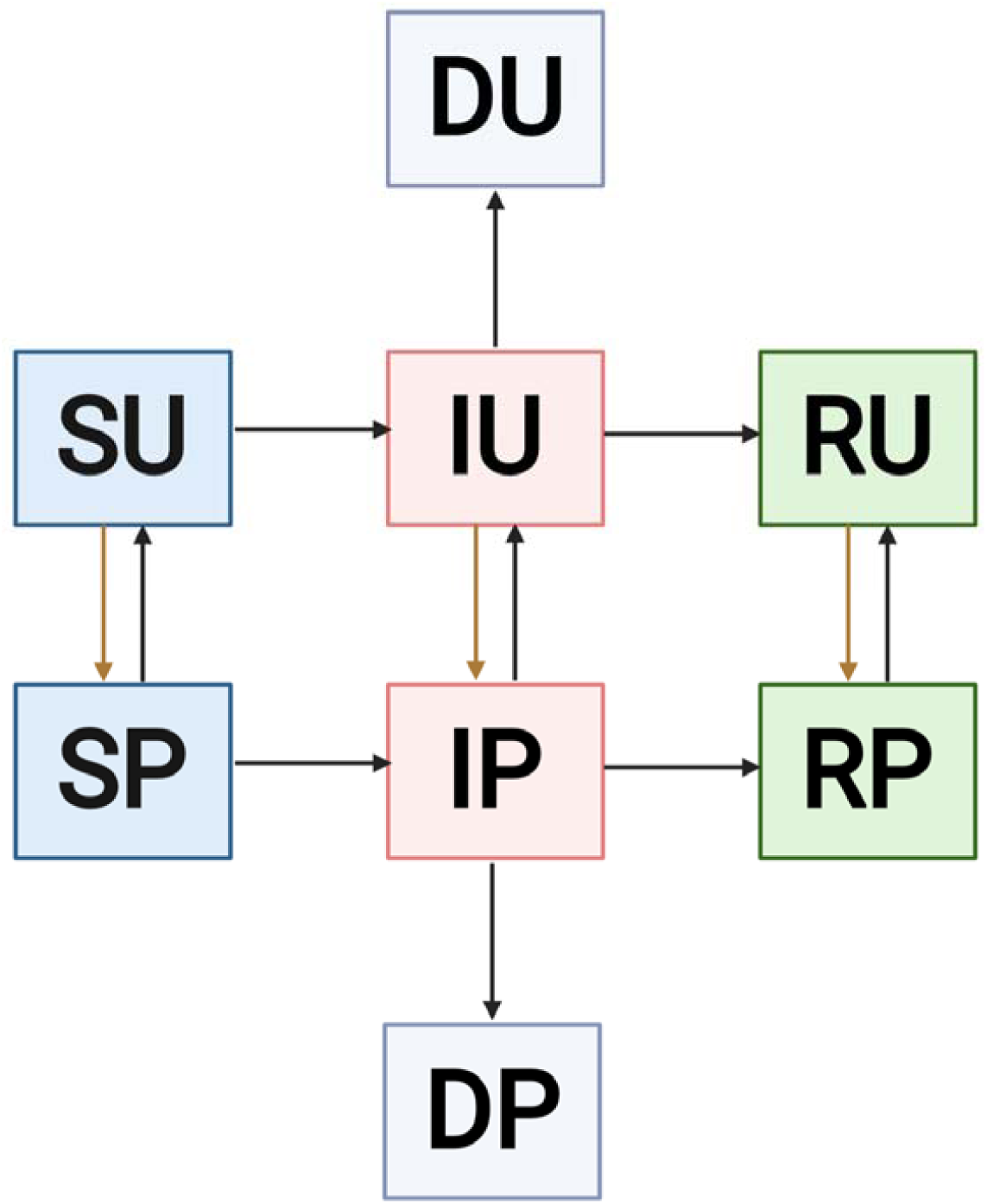
Compartmental diagram for non-pharmaceutical intervention model that tracks status with respect to infection and attitude toward protective behaviors. The first letter of each compartment name gives the state with respect to the disease transmission process (S=Susceptible, I=Infectious, R=Recovered, D=Deceased) and the second letter of each compartment name gives state with respect to awareness-driven protective behavior (U=Unprotective, P=Protective). Squares are colored based on state with respect to disease. Potential transitions are indicated with arrows. Brown arrows indicate awareness-based adoption of protective measures. This diagram corresponds to the model described in Equation 1.

**Supplementary Figure 2.**
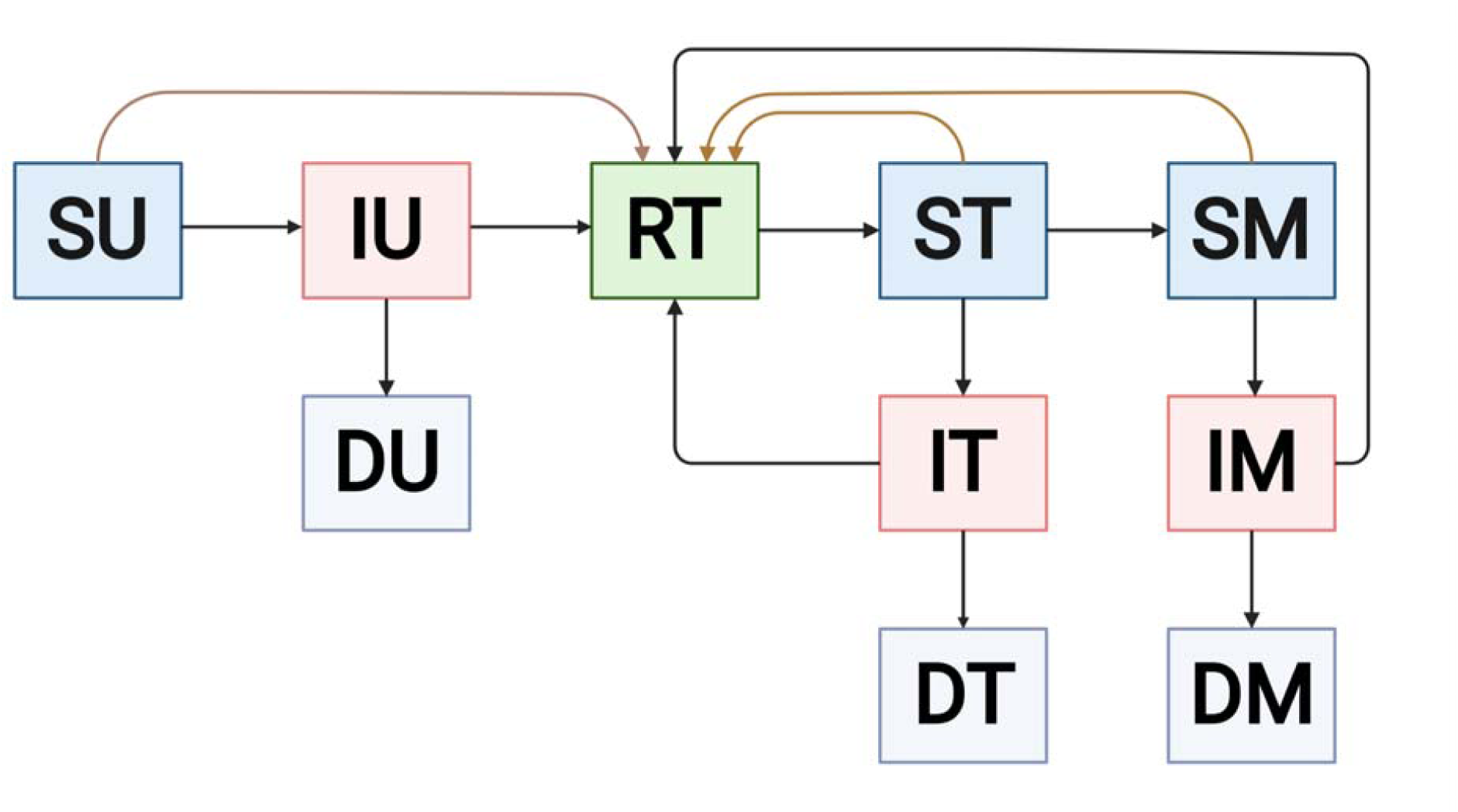
Compartmental diagram for vaccination model that tracks status with respect to infection and immune status. The first letter of each compartment name gives the state with respect to the disease transmission process (S=Susceptible, I=Infectious, R=Recovered, D=Deceased) and the second letter of each compartment name gives immune status (U=Unprotective, T=Transmission and Mortality-Blocking, M=Mortality-Blocking alone). Squares are colored based on state with respect to disease. Potential transitions are indicated with arrows. Brown arrows indicate awareness-based vaccination. This diagram corresponds to the model described in Equation 3.

### Effects of awareness separation on protective behavior and infections

To understand the mechanism by which awareness separation reduces between-group differences in Figure 1, we consider early disease and awareness dynamics for both groups given separated mixing (*ϵ* = 0.99) and uniform or separated awareness (*ϵ* = 0.99and *ϵ* = 0.99respectively).

**Supplementary Figure 3.**
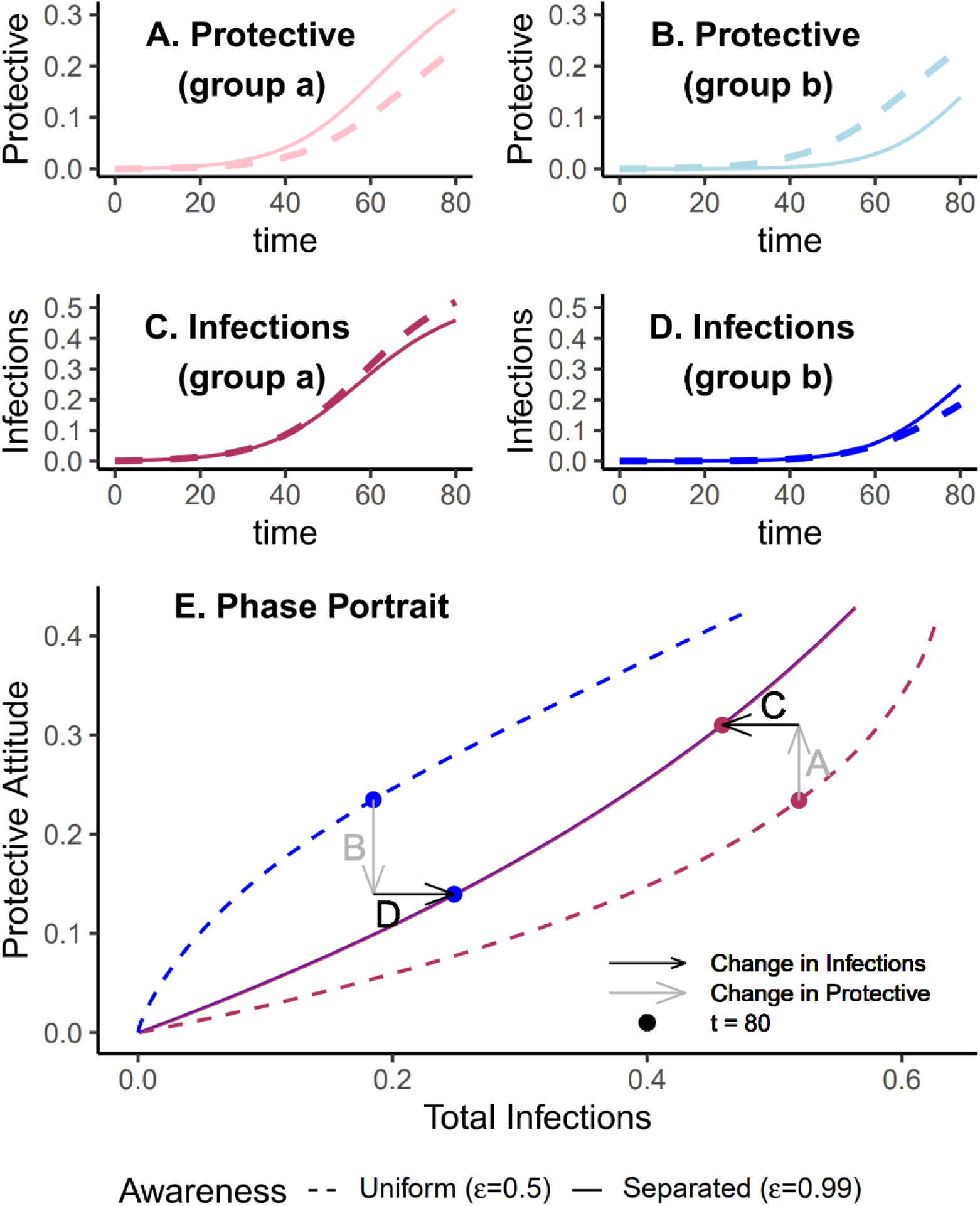
Separated awareness reduces between-group differences by removing group b’s awareness of the emerging epidemic and augmenting group a’s response to the introduction of the pathogen. We initialize our model using the same parameters as Figure 1 with separated mixing (*h* = 0.99). We compare uniform awareness (*ϵ* = 0.5; dashed lines) and separated awareness (*ϵ* = 0.99; solid lines). At the top, we compare early time series (through *t* = 80) of (A) protective attitude prevalence in group a; (B) protective attitude prevalence in group b; (C) infection prevalence in group a; (D) infection prevalence in group b. Panel E is a phase portrait of protective attitude prevalence against total infections in group a (maroon) and group b (blue). Points indicate values at *t* = 80, corresponding to the end of the time series in panels A-infections (black) at *t* = 80 for separated versus uniform awareness, with letters D. Arrows indicate differences in protective attitude prevalence (gray) and total corresponding to time series panel labels.

### Awareness separation reduces effects on mortality of different between-group differences

We demonstrate that the finding in Figure 1 applies across alternative scenarios where the pathogen is introduced in both groups at the same prevalence, but the groups differ in their transmission coefficients (*β*), infection fatality rates (*µ*), or infectious period 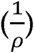 (Supplementary Figures 4, 5, 6). Note that, when transmission coefficient (*β*) varies between groups, contacts between group *a* and group *b* will have transmission Coefficient 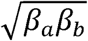, the geometric mean of the transmission coefficient of both groups.

Differences between groups that directly influence force of infection such as variation in transmission coefficient and infectious period, lead to differences in epidemic shape between the groups when mixing is separated (Supplementary Figures 4, 5). Given uniform awareness, epidemic shape is unaffected by mixing separation when group differences do not directly affect the transmission process (e.g., heterogeneity in infection fatality rates (Supplementary Figure 6). In all scenarios, separated awareness decreases differences in deaths between the two groups, although it may not eliminate differences in epidemic burden. In scenarios where groups have different forces of infections, differences in infections are also reduced with separated awareness (Supplementary Figures 4, 5). However, separated awareness increases the difference in infections when groups have different infection fatality rates, as observed in the vaccination scenario in the main text (Figures 3, 4).

**Supplementary Figure 4.**
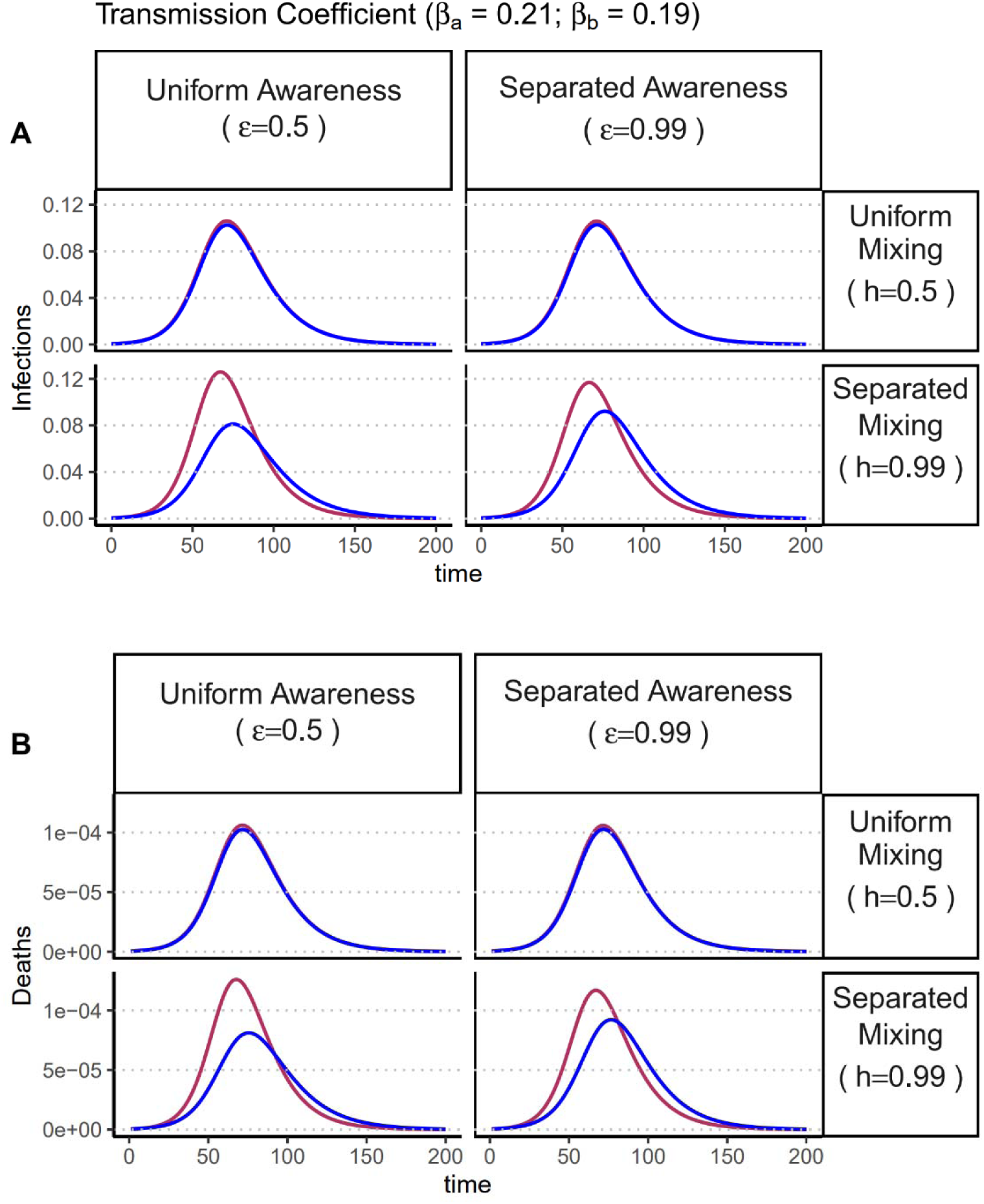
Separated awareness reduces differences in epidemic size between groups in epidemic size that arise from differences in transmission rates coupled with separated mixing. Plots of (A) infections and (B) deaths over time in group a (maroon) and group b (blue). We consider different levels of awareness separation [left column: uniform awareness (*ϵ* = 0.5); right column: separated awareness (*ϵ* = 0.99)] and mixing separation [top row: uniform mixing (*h* = 0.5); bottom row: separated mixing (*ϵ* = 0.99)]. The groups are initialized so that group a has a greater transmission coefficient than group *b* (*β*_*a*_ = 0.21 and *β*_*b*_ = 0.19). We assume the pathogen is introduced in both groups at prevalence 0.0005. All other parameter values are the same as those used in Figure 1: infectious period 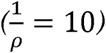, infection fatality rate (*µ*= 0.01), protective measure efficacy (*k* = 0.3), responsiveness (*θ* = 100), memory (*ℓ*= 1), and fatigue (*ν=* 0).

**Supplementary Figure 5.**
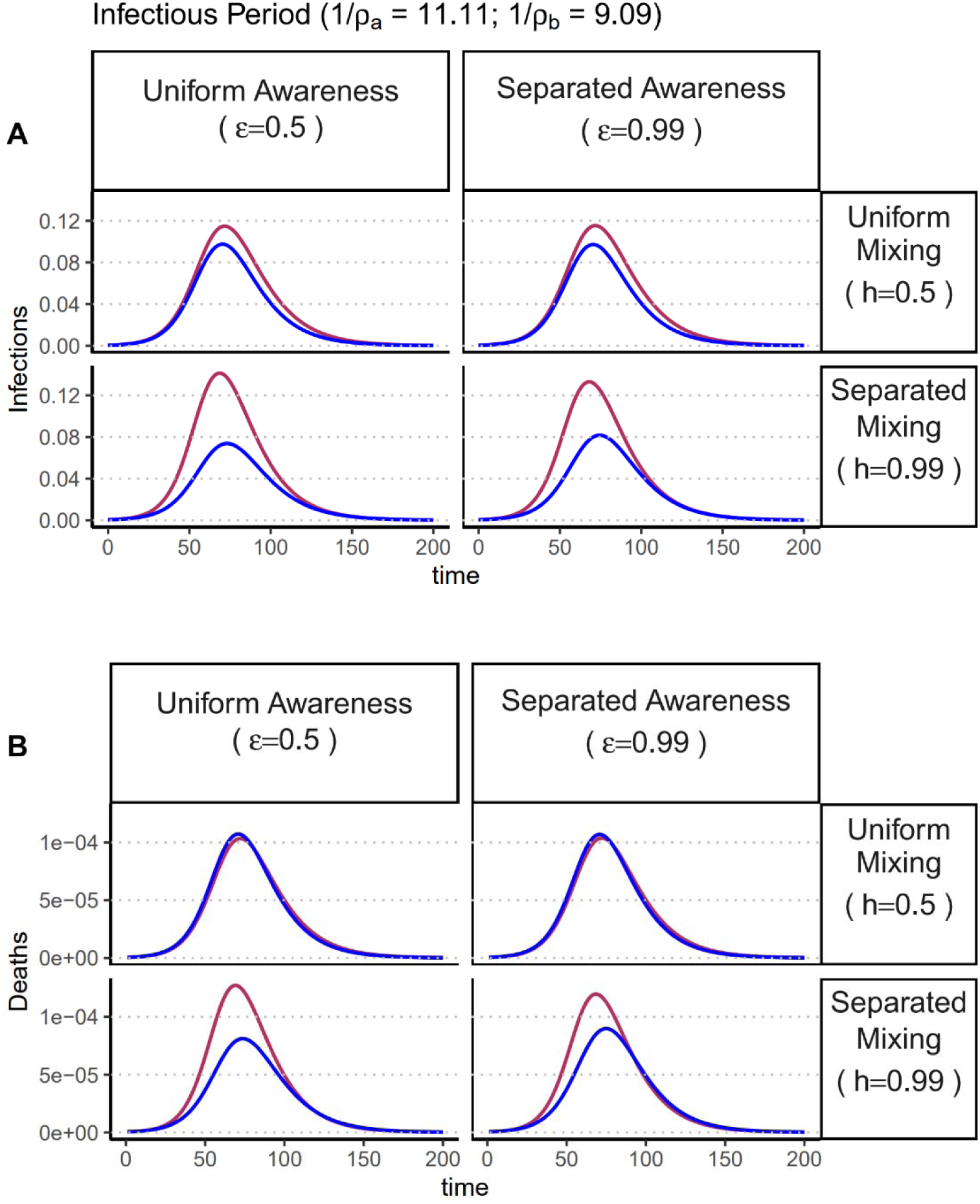
Separated awareness reduces differences in epidemic size between groups in epidemic size that arise from differences in infectious period coupled with separated mixing. Plots of (A) infections and (B) deaths over time in group a (maroon) and group b (blue). We consider different levels of awareness separation [left column: uniform awareness (*h* = 0.5); right column: separated awareness (*ϵ* = 0.99)] and mixing separation [top row: uniform mixing (*h* = 0.5); bottom row: separated mixing (*h* = 0.99)]. The groups are initialized so that group a has a longer infectious period than group *b*(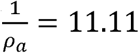 and 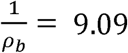). We assume the pathogen is introduced in both groups at prevalence 0.0005. All other parameter values are the same as those used in Figure 1: transmission coefficient (*β*), infection fatality rate (*µ* = 0.01), protective measure efficacy (*k*= 0.3), responsiveness (*θ* = 100), memory (*ℓ*= 1), and fatigue (*ϕ*= 0).

**Supplementary Figure 6.**
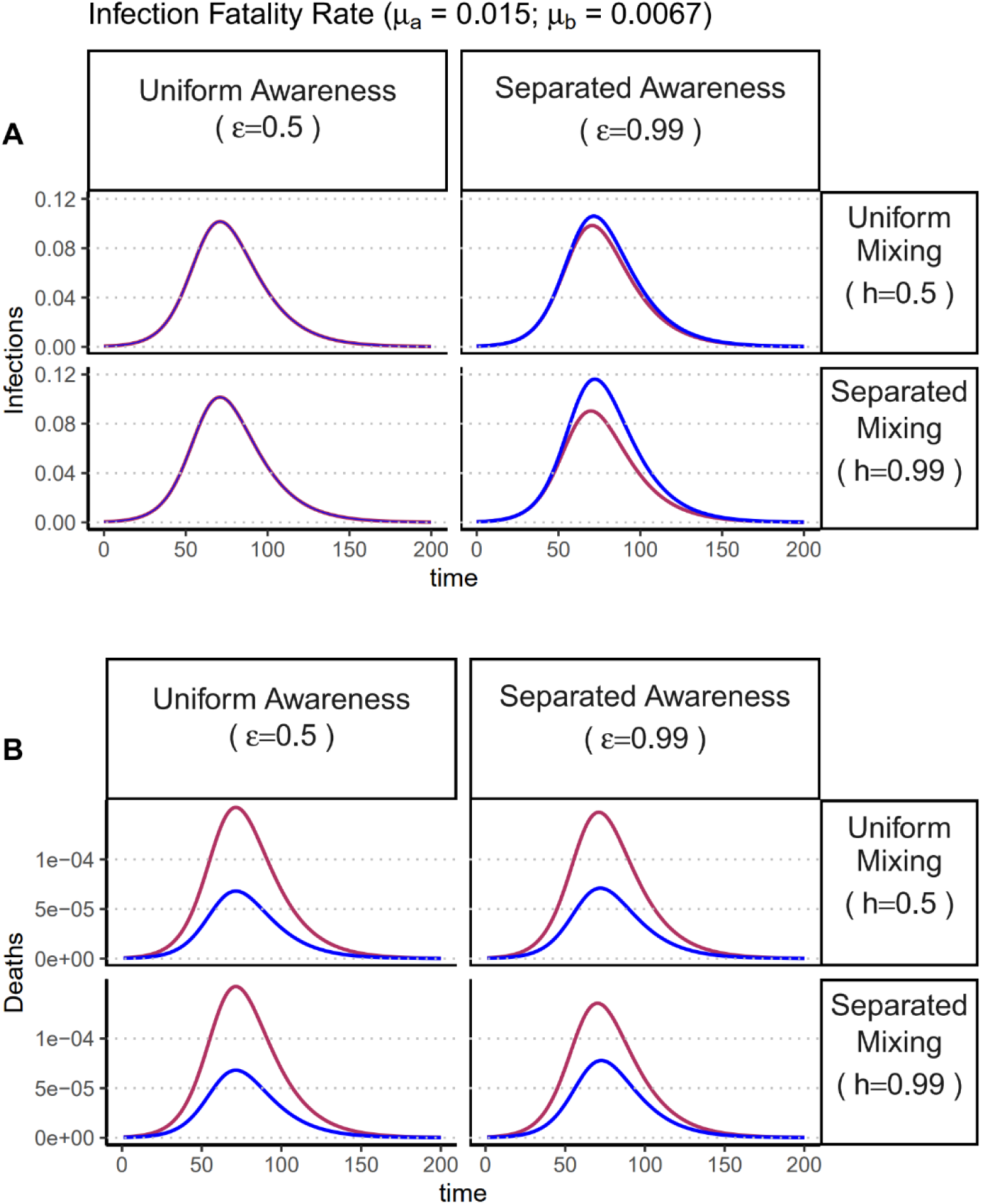
Separated awareness reduces differences in mortality between groups arising from differences in their infection fatality rates and causes differences in infections between the groups. Plots of (A) infections and (B) deaths over time in group a (maroon) and group b (blue). We consider different levels of awareness separation [left column: uniform awareness (*ϵ* = 0.5); right column: separated awareness (*ϵ* = 0.99)] and mixing separation [top row: uniform mixing (*h* = 0.5); bottom row: separated mixing (*h* = 0.99)]. The groups are initialized so that group a has a higher infection fatality rate than group b (*µ*_a_ = 0.015 and *µ* = 0.0067). We assume the pathogen is introduced in both groups at prevalence 0.0005. All other parameter values are the same as those used in Figure 1: transmission coefficient (*β*), infectious period 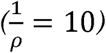, protective measure efficacy *k* = 0.3), responsiveness (*θ* = 100), memory (*ℓ*_ 1), and fatigue (*ν*= 0).

